# Towards Monitoring of Global Health Research: An Exploratory Analysis of Transparency and Stakeholder Engagement

**DOI:** 10.1101/2024.12.30.24319762

**Authors:** Samruddhi Yerunkar, Nicole Hildebrand, Daniel Strech

## Abstract

**Introduction:** Global health research (GHR) requires transparent practices and stakeholder engagement to maximize impact. While monitoring systems exist for clinical research transparency in high-income countries, there is limited systematic assessment of these practices in global health research. This study evaluated methods for monitoring GHR transparency and engagement practices using established indicators.

**Methods:** We analyzed three samples: (1) 200 interventional trials from ClinicalTrials.gov (2008-2019) focused on tuberculosis and maternal health, with two-thirds from low-and-middle-income countries; (2) 200 trial publications from global health journals (2011-2023); and (3) research outputs from global health funder websites. We assessed registration timing, result reporting, open access status, and stakeholder engagement using standardized indicators.

**Results:** Among registry trials, 37% were prospectively registered, 65% published results in journals, and 15% reported summary results in ClinicalTrials.gov. Only 34% reported results in any format within 24 months of completion. For journal publications, 72% were freely accessible, and among a subsample of 100 articles, 23% included stakeholder engagement statements. The funder website sample yielded insufficient metadata for systematic analysis.

**Conclusion:** Our findings demonstrate that monitoring GHR transparency and engagement is feasible using trial registries and journal publications, though funder websites currently lack adequate tracking data. While open access rates are encouraging, timely result reporting and stakeholder engagement documentation need improvement. These results highlight opportunities for developing GHR-specific monitoring approaches through collaborative efforts among global stakeholders.

**Funding:** Berlin University Alliance

**What is already known on this topic:** Previous studies have highlighted gaps in transparency and patient engagement in clinical trials. To address these, various tools, such as [national](https://frenchopensciencemonitor.esr.gouv.fr/) and [university](https://quest-dashboard.charite.de/#tabStart) dashboards, have been developed to monitor practices. In global health research (GHR), this monitoring has been primarily led by intergovernmental agencies; however, academic research can complement these efforts by enhancing rigor and driving actionable change. This study proposes a methodological approach to identify global health studies for analysis with established research indicators and aims to suggest complementary strategies for analyzing GHR.

**What this study adds:** This study introduces a methodology suitable for assessing GHR studies with transparency and engagement indicators. We sourced data from trial registry, global health journals, and funder websites, providing accompanying code and data to facilitate reproducibility for other researchers in the field. Unlike prior analyses in global health context that often emphasize LMICs, our approach incorporates both LMIC and HIC studies, recognizing the collaborative foundation of GHR and encouraging shared contextualized findings. Our findings offer empirical data while underscoring the need for expanded collaboration to refine indicators and methodologies, enhancing the contextual relevance of GHR monitoring.

**How this study might affect research, practice or policy:** Collaborative efforts among researchers, funders, and communities are essential to develop new methodologies and indicators that focus on actionable change highlighted by continuous monitoring. While often led by large organizations, GHR monitoring can be enriched by academic research, making it more contextualized and accessible to global health researchers. By proposing initial methodologies, we provide a foundation that can be refined along with indicators that can be contextualized, both of which are essential for driving effective global health research.

## 1. Introduction

Global health research (GHR) is a collaborative, transnational research approach aimed at promoting health equity for all (1). The field addresses major health challenges that affect large populations worldwide, often focusing on severe conditions with substantial impact on mortality and quality of life, particularly in resource-limited settings. Recent surges in GHR activities, especially spurred by the COVID-19 pandemic, have highlighted both the field’s potential and its challenges in delivering timely evidence for decision-making.

Research transparency and community engagement are crucial elements across all fields of health research. In GHR, these practices take on additional dimensions due to the field’s unique characteristics. For instance, many global health conditions affect substantial populations across different regions and cultural contexts, making rapid knowledge translation particularly important. Limited research infrastructure in many settings and historical power imbalances between researchers and communities further emphasize the need for transparent research practices and meaningful stakeholder engagement.

The timely dissemination of research findings through transparent practices such as prospective registration, comprehensive results reporting, and open-access publication is essential for evidence-based healthcare globally. In GHR, barriers to accessing research findings can be especially problematic - paywalled publications may be inaccessible to healthcare providers, researchers, and patients in low-resource settings, potentially widening rather than reducing health inequities. Similarly, the transnational and cross-cultural nature of GHR makes genuine patient and stakeholder engagement (PSE) particularly relevant for ensuring research relevance, cultural appropriateness, and eventual implementation.

Historically, global health monitoring was primarily led by major inter-governmental agencies. However, academic research has increasingly complemented these efforts, enhancing the methods and rigour of global health measurement to support better decision-making by agencies, governments, and donors (2). While significant efforts have been made to monitor trial transparency and engagement in Europe and North America (3-7), gaps remain in assessing these practices on a global scale (8). For instance, the EU TrialsTrackers (http://eu.trialstracker.net/) specifically monitor whether summary results are posted on the EU clinical trials register and national-level dashboards have also been developed, providing a detailed breakdown of practices and helping stakeholders identify and address areas needing improvement (7, 9). Similar GHR monitors can help assess the current landscape, highlight actionable areas, and guide stakeholders—including patients, researchers, physicians, and global leaders—in making informed decisions.

Improved global health monitoring further requires new technologies and methods, as well as established norms and standards to facilitate global reporting (10). To build an effective monitoring system for global health, it is important to first define what constitutes global health studies and then assess with appropriate indicators. We explored three methods for generating a sample of global health studies: (1) clinical trial registries focusing on global health conditions or diseases affecting populations globally, (2) publications from global health journals, and (3) research outputs mentioned on global health funders’ websites. Using indicators established in previous studies, we assessed a sample of these studies to provide empirical data. We aim to demonstrate how past methodologies for monitoring clinical research can be applied to GHR, highlighting opportunities and challenges to enhance the implementation of monitoring systems in the GHR.

## 2. Methods

### 2.1. Sampling Strategy

We explored three complementary approaches to identify global health research studies: (1) clinical trial registries, using selected global health conditions as proxy, (2) publications from journals with an explicit global health focus, and (3) research outputs listed on global health funder websites. Each source offered distinct advantages and challenges for identifying and tracking global health studies. Below, we describe the methodology and purpose.

#### 2.1.1 Clinical Trial Registry with a disease-based approach

We systematically searched trial registry of ClinicalTrials.gov for interventional trials with start date between January 1, 2008 to completion date March 31, 2019. This eligibility requirement was based on the mandate for clinical trial registration in registries under both US laws and journal policies following ICMJE guidelines (11) passed on January 1, 2008. For our analysis, we focused on two health conditions known to have significant global burdens: maternal health issues (including postpartum depression, maternal sepsis, and maternal anaemia) and tuberculosis. These conditions were chosen based on their significant global health burden and their relevance to our ongoing larger research project, where one of our team members has a specific interest in maternal health, particularly postpartum conditions. We excluded international multicentric trials, observational studies, and those with missing location or site information, as well as withdrawn studies. Trial locations were categorized as high-income countries (HIC) or low-to-middle-income countries (LMIC) according to the World Bank country classification (12). For our exploratory analysis, we randomly selected 100 trials per condition. To address the imbalance, with more than half of the trials originating from HICs, we ensured that two-thirds of the selected trials were from LMICs. This approach ensured equitable representation and also addressed the historical disparity in research quality assessments between HICs and LMICs. In this sample, we aim to demonstrate the feasibility of evaluating registration practices and the timeliness of result reporting by examining both summary results and publications from a sample of clinical trials within a global health context.

To locate trial publications, two independent reviewers (NH and SSY), followed a predefined search manual to identify the earliest publications of trial results (see [search manual](https://osf.io/9hktj)). Searches were conducted in both the registry and Google, using trial IDs and relevant terms. Eligible publications included peer-reviewed articles or preprints with over 500 words, matched to the trial by design, population, intervention, and comparator (if applicable). Additionally, the publication’s primary outcome measure (or first mentioned outcome) had to be listed as an outcome measure in the registration, whether classified as primary or not.

#### 2.1.2 Global Health Journals

Our second approach focussed on generating a sample of global health studies from peer-reviewed journals specializing in global health. We identified 20 journals that either had ‘global health’ or ‘international health’ in their title (see [list of journals](https://osf.io/a6mtq)). Using the Cochrane Highly Sensitive Search Strategy (Box 3b strategy), we located randomized trials published after January 1^st^, 2011, in PubMed and cross-referenced these results with our selected journals. Including trial publications post-2011 allowed a three-year window for dissemination of results, following the ICMJE mandate for prospective trial registration, effective from 2008. Our search yielded 1,125 records of trials published after 2011. An author (SSY) screened a random sample (n = 300), categorizing them as primary reports of trial results or secondary analyses by the original or different trial investigators. For analysis, we included the first 200 primary reports and secondary analyses by same investigators (see Supplementary Figure 1 flowchart). This sample was used to evaluate how trial results are reported in GHR, examining adherence inclusion of data sharing, and community engagement statements.

#### 2.1.3 Global Health Funder website

The third method aimed to explore the use of global health funder websites as potential sources for generating global health studies. We investigated funder websites to determine if they provided sufficient meta-data, such as trial registration numbers and registry links, to allow for the tracking of funded clinical trials and to assess transparency. This involved searching World RePORT (13), a database for tracking funding data for some largest biomedical funders, as well as a list of global health funders compiled by colleagues for a prior project. Our search revealed 20 global health funders and 1 of them had explicit GH programs with available funding meta-data, sufficient to track funded clinical trials (see [list of global health funders](https://osf.io/acm4e)).

We focused on the Fogarty International Center (FIC) at the National Institutes of Health (NIH), which supports global health research. Using the NIH RePORTER tool, we identified a list of 73 clinical trials and 9515 publications supported by 331 core projects. However, after applying the inclusion criterion with regard to date, the number of relevant trials was very limited, leading us to exclude this sample from our analysis.

### 2.2. Software

We used the aactr R package (14) to assess registration and summary result reporting in registry of trials. The R packages oddpub (15) and roadoi (16) were used to assess availability statements and open access status respectively in trial publications. Map was created using the Free and Open Source QGIS (17). Data cleaning steps and statistical analyses were performed using R (18). All analysis scripts are available under an open license on GitHub https://github.com/quest-bih/prodigy.

## 3. Results

This section provides an independent analysis of two samples: the first focuses on interventional trials from ClinicalTrials.gov, while the second examines trial result publications from global health journals. Data from the third funder website sample was limited and not analyzed.

### 3.1 Follow-up of clinical trials from Clinical trial registry

We included data from 200 interventional trials (2009 – 2019) with 2/3^rd^ representation from LMIC, enrolling 115,929 participants. Dominant trial characteristics were small sample size (<100) and open labelled (Table 1). The most common trial locations were the United States (n = 25), South Africa (n = 24), and China (n = 23) (Figure 1).

**Table 1.** Established indicators used for assessing transparency and stakeholder engagement in global health research samples.

| Indicator | Details | Assessment | Impact |
| --- | --- | --- | --- |
| **Follow-up of clinical trials from clinical trial registry with a disease-based approach** |  |  |  |
| Time to registration | Before any clinical trial is initiated its details must be registered in a publicly available, free-to-access, searchable clinical trial registry complying with WHO's international agreed standards<sup>1</sup>. | Automated check by comparing the trial registration date with the trial start date. We report on<br>1) before the trial starts (prospective registration),<br>2) within 60 days after the trial starts (early registration),<br>3) after 60 days (late registration) | Transparency, unbiased reporting |
| Result reporting | Results can be reported in the following way: Published articles in a peer-reviewed journal (24 months from trial completion); Trial results uploaded in the trial registry (12 months from trial completion)<sup>1</sup>. | Automated check for summary result reporting in the registry and manual search<sup>2</sup> for published articles. We report on<br>1) Summary result (SR) in registry<br>2) Journal publication<br>3) SR or journal publication within 1 year after trial completion<br>4) SR or journal publication within 2 years after trial completion<br>5) SR or journal publication within 4 years after trial completion | Transparency, informed decision making |
| Open Access | Publications describing clinical trial results should be open-access<sup>1</sup>. | Automated check for publication status (open access) for manually searching research publications of clinical trials. | Transparency, accessibility |
| **Analysis of clinical trial result publication in global health journals** |  |  |  |
| Community Engagement/Patient Stakeholder Reporting | Involvement of patients and other relevant stakeholders (such as caregivers, healthcare providers, patient advocacy groups, policymakers, and the community) in any phase of the clinical study process. | Manual review of publications for stakeholder involvement statements. | Practical relevance, inclusivity |
| Open Access | Publications describing clinical trial results should be open-access<sup>1</sup>. | Automated check for publication status (open access) | Transparency, accessibility |
| Data sharing statement | Data sharing, archiving, depositions, and accessibility of data in journal articles with or without request | Automated check for declarations on data sharing and availability. | Transparency, reproducibility |
| Code sharing statement | Support sharing of health research datasets whenever appropriate | Automated check for open code statements. | Transparency, reproducibility |

<sup>1</sup> Text referenced from WHO Joint statement on public disclosure of results from clinical trials (https://www.who.int/news/item/18-05-2017-joint-statement-on-registration).
<sup>2</sup> Manual searches are done according to the search manual for locating result publications (https://osf.io/9hktj).

**Table 2.** Characteristics of included clinical trials.

| Trial characteristic<sup>1</sup> | Tuberculosis (n =100) | Maternal Health Conditions (n = 100) | All trials, N= 200 |
| --- | --- | --- | --- |
| **Recruitment status** |  |  |  |
| Completed | 79 (79) | 76 (76) | 155 (77.5) |
| Terminated | 2 (2) | 8 (8) | 10 (5) |
| Unknown status | 19 (19) | 16 (16) | 35 (17.5) |
| **Enrolment** |  |  |  |
| 1- 100 | 49 (49) | 40 (40) | 89 (44.5) |
| 100 - 500 | 32 (32) | 44 (44) | 76 (38) |
| >500 | 19 (19) | 16 (16) | 35 (17.5) |
| **Masking** (missing, n =1) |  |  |  |
| Open label | 62 (62) | 53 (53) | 115 (57.5) |
| Any masking | 37 (37) | 47 (47) | 84 (42) |
| **Design** (missing, n =29) |  |  |  |
| Non-randomized | 10 (10) | 15 (15) | 25 (12.5) |
| Randomized | 76 (76) | 70 (70) | 146 (73) |
| **Phase** |  |  |  |
| I | 19 (19) | 5 (5) | 24 (12) |
| I-II | 3 (3) | 1 (1) | 4 (2) |
| II | 21 (21) | 14 (14) | 35 (17.5) |
| II-III | 2 (2) | 2 (2) | 4 (2) |
| III | 9 (9) | 13 (13) | 22 (11) |
| IV | 11 (11) | 24 (24) | 35 (17.5) |
| Not applicable | 35 (35) | 41 (41) | 76 (38) |

<sup>1</sup>Total count may differ between categories because of missing data (number missing shown if >0).

#### Trial registration and result reporting

Of the 200 trials, 74 (37%) were registered prospectively. Regarding result reporting, from all 200 trials that were completed until 2019 or earlier and thus had 4 or more years to report results, 129 trials (65%) had results published in journals, and 30 (15%) reported summary results in registries. The number of trials that reported any results (either via journals or summary results) within one, two, or four years after completion was 29 trials (15%), 67 trials (34%) and 116 (58%) (Table 3).

**Table 3.**
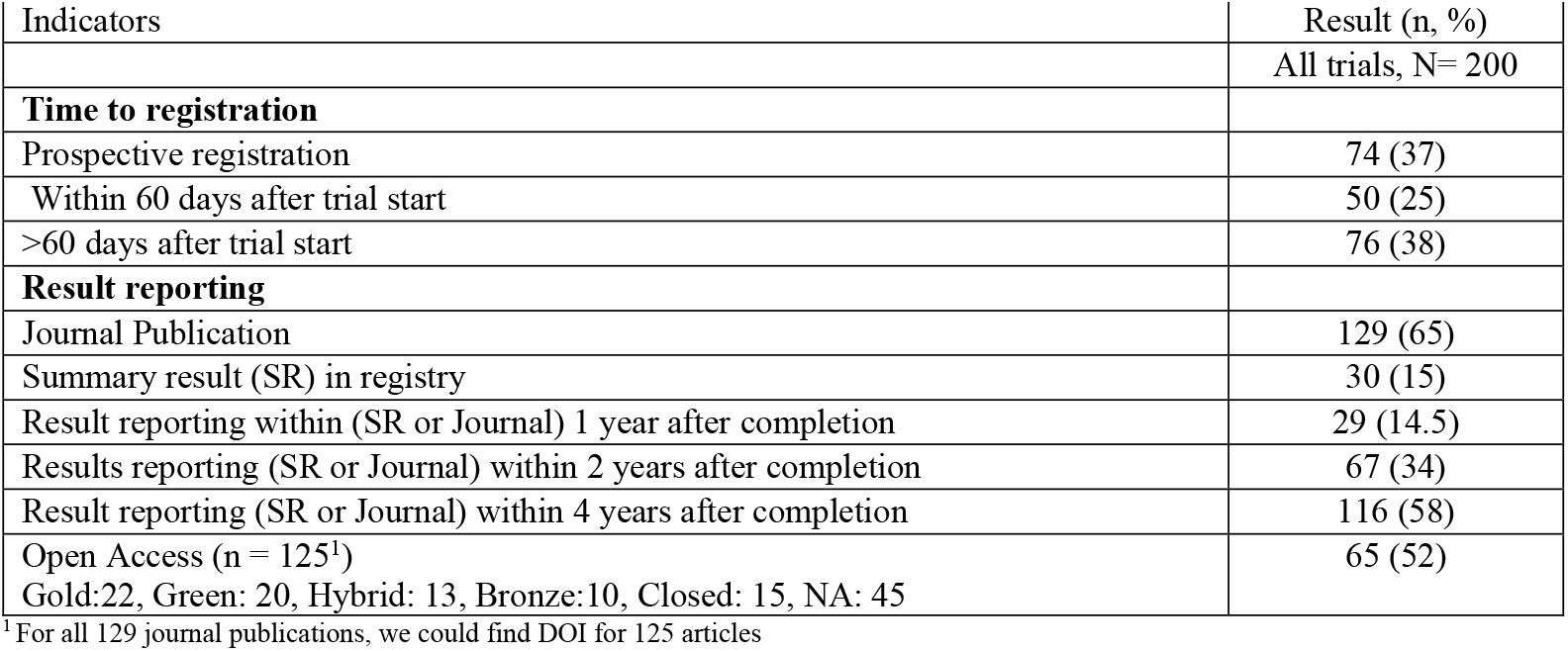
Registration and result reporting of included clinical trials.

For journal result publications identified using a search manual, the reviewer independently located relevant publications with an interrater reliability of 83%. When different publications were selected, 100% agreement was reached on selecting the earliest matching publication. All matched publications were peer-reviewed articles, with majority – specifically, 60% (78 /129) - linked in the registry.

### 3.2 Analysis of trial results in publications from GH journals

Of the 200 trial publications in global health journals between 2011 to 2023, 72% (144) were Gold Open Access (freely accessible), 22.5% (45) were Hybrid Open Access (partially accessible), and 1.5% (3) were closed access. We were able to download 127 of the 200 PDFs to assess data and code availability. Among these, 59% (75) included a data-sharing statement, often specifying availability upon request, and only 0.8% (1) included a code-sharing statement. A random sample of 100 articles was reviewed for stakeholder engagement statements using a predefined protocol (see [search manual](https://osf.io/g459j)) with 23% (23) reporting relevant content (see [extracted statements](https://osf.io/knf23)).

**Table 2:** Transparency and community engagement in included trial result publication.

| Indicators | Result (n, %) |
| --- | --- |
| Open Access (n = 200) | 197 (99) |
| Gold: 144, Green: 8, Hybrid:45, Closed:3, |  |
| Data sharing statement including ‘available upon request’ (n = 127<sup>1</sup>) | 75 (59) |
| Code sharing statement (n = 127<sup>1</sup>) | 1 (0.8) |
| Community/Stakeholder engagement statement (n = 100<sup>2</sup>) | 23 (23) |

<sup>1</sup>For all 200 we had 127 PDFs, <sup>2</sup>Checked for a random sample of 100 publications

## 4. Discussion

This study aimed to explore approaches for monitoring transparency and patient/stakeholder engagement in Global Health Research (GHR) while providing empirical data on the current state of these practices. We tested three different sampling strategies and found that trial registries focusing on global health conditions and global health journals provide viable data sources for monitoring, while funder websites currently lack sufficient metadata for systematic tracking.

Among 200 registered trials, 37% were prospectively registered, and 65% had published results in journals, though only 15% reported summary results. Within 24 months of trial completion, only 34% reported their result in any format and thus demonstrated timely results reporting. These findings on timely results reporting are comparable to previous studies examining clinical trial transparency in other contexts, such as German university medical centers (43%) (6), Nordic countries (52%) (5), and US academic medical centers (36%) (19). Patient/Stakeholder engagement was mentioned in 23% of publications, providing a baseline for more detailed future analyses of engagement practices in GHR.

Our methodological approach had several strengths and limitations. The disease-based sampling strategy using trial registries proved effective for identifying GHR studies but required careful selection of conditions. Among identified publications, 60% were properly linked to their registry entries, indicating moderate but improvable compliance with WHO recommendations for result traceability. The journal-based approach successfully identified GHR publications but is limited by the need to rely on “global health journals” to represent global health research. The funder-based approach highlighted current gaps in tracking systems that need to be addressed to enable comprehensive monitoring. A key methodological challenge was ensuring balanced representation between high- and low-income countries. Our approach of oversampling LMIC-based studies (2/3 of trial registry sample) provides one potential solution, though optimal sampling strategies for GHR monitoring should be further discussed with stakeholders.

While the overall rates of timely reporting are similar to other contexts, timely results reporting is particularly important in GHR given that many global health conditions represent a high global burden of disease (GBD). Although it would be problematic to claim that timely reporting is more important in some research fields than others, the often substantial disease burden addressed in GHR emphasizes the need for rapid knowledge translation to inform evidence-based healthcare decisions. The observed high rate of open-access publications (99%) in our sample is particularly encouraging, as paywalled research results would disproportionally affect healthcare providers and researchers in low-resource settings. This rate markedly exceeds the average of 28% open access found across scholarly literature. While this high rate may partly reflect our sampling of global health journals with established open-access policies and increasing pressure on global health researchers to publish transparently, it nevertheless demonstrates substantial progress in making GHR findings widely accessible.

The increasing adoption of institutional monitoring tools, as demonstrated by Franzen et al. (7), could help track and improve transparency in GHR. Such tools should incorporate core open science practices recently identified through expert consensus (20) while considering GHR-specific needs. The baseline measurement of patient/stakeholder engagement reporting provides important insights for future development. Given that GHR often involves transnational and cross-cultural research contexts, meaningful engagement of local communities and stakeholders is particularly crucial. However, our finding that only 23% of publications mentioned engagement activities suggests room for improvement. As demonstrated by Weschke et al. (21), more detailed analyses of engagement reporting can reveal important insights about the quality and impact of these practices.

In our analysis, we assessed only a sample of studies to provide empirical data on how GHR can be monitored. However, global health research is inherently collaborative, requiring active input from researchers and stakeholders. Future efforts should identify additional indicators for monitoring, address challenges in informing researchers about their performance in transparency and engagement, and expand on our findings using new samples and methodologies. Continued monitoring can drive improved practices, fostering more transparent and inclusive GHR to support evidence-based global health advancements.

## Data Availability

All underlying raw and processed datasets for this study are available for download from the Open Science Framework (https://osf.io/hqdns/).

## 5. Data sharing

All the code for this study is available via open-source licensing on GitHub (https://github.com/quest-bih/prodigy). All underlying raw and processed datasets for this study are available for download from the Open Science Framework (https://osf.io/hqdns/).

## 6. Declaration of interests

We declare no competing interests.

## 7. Acknowledgments

This study is part of the [PRODIGY project](https://www.berlin-university-alliance.de/foerderung-angebote/projects/Projektliste_INT/prodigy-practices-of-development.html) and we would like to thank Malek Bajbouj, Isabel Dziobek, Eric Hahn, Thi Minh Tam Ta, María Jose Lobeda-Garzón as well as Sarah Weschke, Vladislav Nachev and Delwen Franzen who contributed their ideas and knowledge to make this study possible.

## 8. Author contributions

Conceptualization: SSY, DS

Formal analysis: SSY

Funding acquisition: DS

Investigation: SSY,

NH Methodology: SSY, DS

Project administration: SSY

Supervision: DS

Writing – original draft: SSY

Writing – review & editing: SSY, NH, DS

## 9. Funding

This work was funded by the Berlin University Alliance (BUA). DS received funding for a work package within the [PRODIGY project](https://www.berlin-university-alliance.de/foerderung-angebote/projects/Projektliste_INT/prodigy-practices-of-development.html). The funder had no role in the study design, data collection and analysis, decision to publish, or preparation of the manuscript.

## Notes

### Competing Interest Statement

The authors have declared no competing interest.

### Summary of Updates

The established indicators for assessing transparency and stakeholder engagement in global health research samples (Table 1) have been updated with a clearer version for improved readability. Spotted typos and misplaced commas have been corrected. The previously non-functional link to the extracted PSE statement has been fixed.

