## Supplementary file 1 for "Towards Monitoring of Global Health Research: An Exploratory Analysis of Transparency and Stakeholder Engagement"

### Supplementary Method details

#### 1. Trial Registry using Global Health Condition

On 13th April 2024, using the ClinicalTrials.gov web front (https://clinicaltrials.gov/), we searched studies by Condition/disease (MeSH terms) in the following way:

| Condition | N |
|---|---|
| Postpartum Depression (also searched for **Depressive Disorders**, **Puerperal**, and **Post Natal Depression**. See Search Details) | 450 |
| Maternal Sepsis (also searched for **Depressive Disorders**, **Puerperal**, and **Post Natal Depression**. See Search Details) | 1403 |
| Maternal Anemia (also searched for **Anemia of pregnancy**, **Pregnant**, **Anemia during pregnancy** and more. See Search Details) | 437 |
| Tuberculosis (See Search Details ) | 1391 |

2. **Global Health Journal**

On 13th April 2024, using the Cochrane Highly Sensitive Search Strategy (Box 3b strategy), we identified randomized trials in PubMed and cross-referenced these results with articles from 20 global health journals.

Box 3. b Cochrane Highly Sensitive Search Strategy for identifying randomized trials in MEDLINE: sensitivity- and precision-maximizing version (2008 revision); PubMed format

**#1 randomized controlled trial [pt]**

**#2 controlled clinical trial [pt]**

**#3 randomized [tiab]**

**#4 placebo [tiab]**

**#5 clinical trials as topic [mesh:noexp]**

**#6 randomly [tiab]**

**#7 trial [ti]**

**#8 #1 OR #2 OR #3 OR #4 OR #5 OR #6 OR #7**

**#9 animals [mh] NOT humans [mh]**

**#10 #8 NOT #9**

PubMed search syntax:
[pt] denotes a Publication Type term;
[tiab] denotes a word in the title or abstract;
[sh] denotes a subheading;
[mh] denotes a Medical Subject Heading (MeSH) term ‘exploded’;
[mesh:noexp] denotes a Medical Subject Heading (MeSH) term not ‘exploded’;
[ti] denotes a word in the title.

Supplementary file 1

**PubMed terms used to extract articles**

((((("The Lancet. Global health"[Journal]) OR ("Journal of global health"[Journal])) OR ("Globalization and health"[Journal])) OR ("Annals of global health"[Journal])) OR ("pathogens and global health"[Journal])) OR ("Global public health"[Journal])) OR ("Global health action"[Journal])) OR ("Global health promotion"[Journal])) OR ("BMJ global health"[Journal])) OR ("Global pediatric health"[Journal])) OR ("Global health, epidemiology and genomics"[Journal])) OR ("Global health research and policy"[Journal])) OR ("The Central African journal of medicine"[Journal])) OR ("Clinical epidemiology and global health"[Journal])) OR ("Global journal of health science"[Journal])) OR ("Global mental health (Cambridge, England)"[Journal])) OR ("international journal of travel medicine and global health"[Journal])) OR ("plos global public health"[Journal])) OR ("Global journal of health science"[Journal])) OR ("journal of global health perspectives"[Journal])) OR ("Global health journal (Amsterdam, Netherlands)"[Journal])) and (((randomized controlled trial[Publication Type]) OR (controlled clinical trial[Publication Type]) OR (randomized[Title/Abstract]) OR (placebo[Title/Abstract]) OR (clinical trials as topic[mesh:noexp]) OR (randomly[Title/Abstract]) OR (randomly[Title/Abstract]) OR (trial[Title])) NOT (animals [mh] NOT humans [mh]))) NOT review [pt]) AND ((english[Filter]) AND (2011:2024[pdat]))) Filters: English, from 2011 – 2024

Note: We additionally excluded articles (n = 9) from “The Central African journal of medicine” journal, which was not explicitly a global health journal. Please see “13-4-2024-global-health-article” for details.

Supplementary file 1

*Supplementary Figure 1 Global Health Journal Analysis Flowchart*

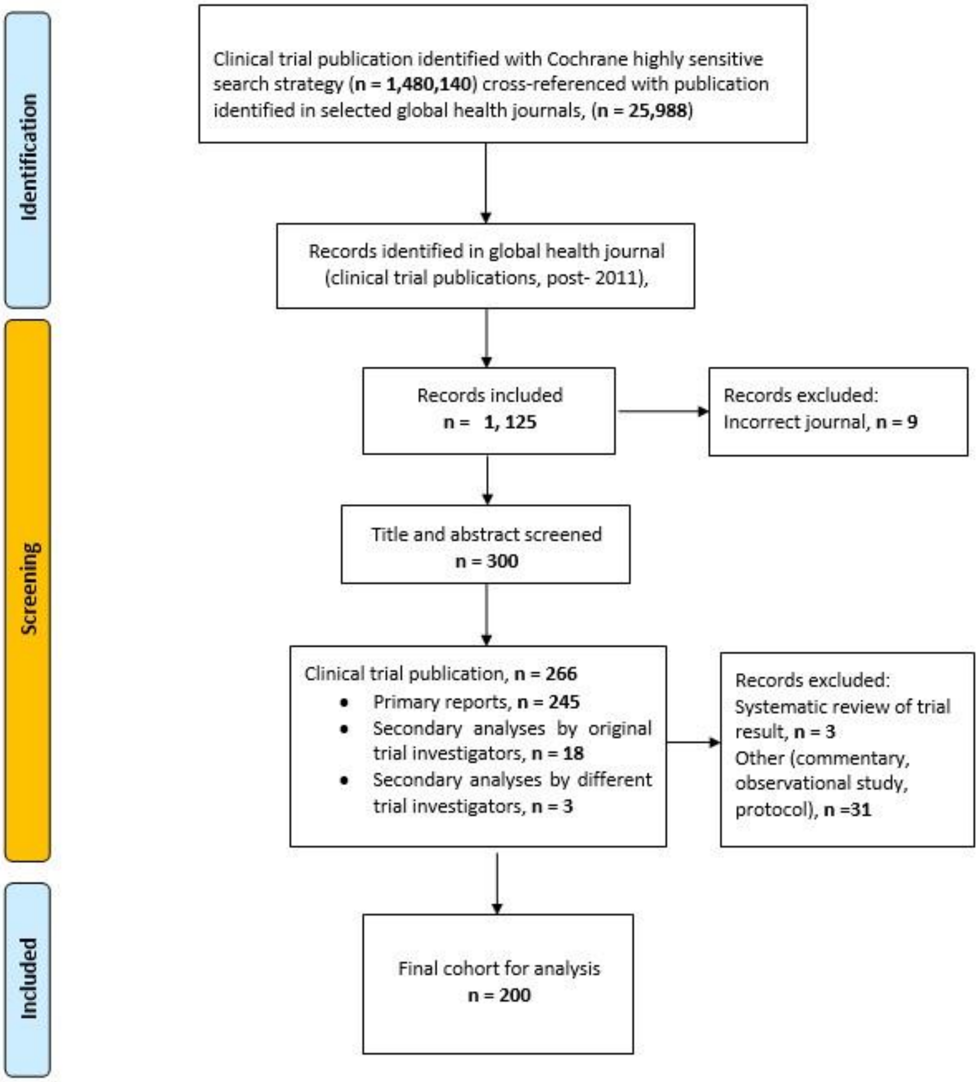

**3. Global Health Funder**

On 13th April 2024, using the NIH RePORTER tool's 'Advanced Project Search' (https://reporter.nih.gov/advanced-search) and selecting the Funding tab, with Agency/Institute/Center set to "John E. Fogarty International Center for Advanced Study in the Health Sciences (FIC)," the search results included:

- 73 clinical trials,
- 9,515 publications,
- Supported by 331 core projects.
